# Continuous respiratory rate monitoring using photoplethysmography technology in patients with Obstructive Sleep Apnea

**DOI:** 10.1101/2023.07.25.23293134

**Authors:** J.M. Gehring, L.C. Saeijs-van Niel, L.P ten Bosch-Paniagua, M.H. Frank

## Abstract

**Purpose:** Respiratory rate is an important physiological parameter whose abnormality could be, in the presence of other suggestive symptoms, supportive of a diagnose of various serious illnesses. Photoplethysmography (PPG) in wearable sensors potentially plays an important role in early disease detection by making respiratory rate measurements more accessible. We investigated the accuracy of a new non-invasive, continuous, wrist-worn and wireless monitoring PPG device (Corsano CardioWatch 287) in measuring respiratory rate (RR) and heart rate (HR) at rest.

**Methods:** Subjects with and without diagnosed Obstructive Sleep Apnea (OSA) underwent simultaneous, continuous PPG and home sleep apnea testing (HSAT) for one night. We assessed the PPG sensor’s measurement accuracy by calculating root-mean-square accuracy (A_rms_) and by performing Bland Altman and correlation analysis. Subgroups were defined based on skin type, hair density, age, BMI, gender and OSA severity.

**Results:** In 26 participants a total of 31083 RR and 38693 HR measurement pairs were obtained. For RR measurements, A_rms_ was 0.60 breaths per minute (brpm). Correlation was high (r = 0.964 (95%CI 0.963 - 0.965)) and 95% Limits of Agreement (LoA) were −1.28 to 1.71 brpm (mean bias −0.14 brpm). For HR measurements, A_rms_ was 0.95 beats per minute (bpm). Correlation was similarly high (r = 0.996 (95%CI 0.996 - 0.996) and 95% LoA were – 2.09 to 1.17 bpm (mean bias −0.46 bpm). Results were comparable across all subgroups, without significant difference in RR accuracy between subjects with and without diagnosed OSA. In free-living conditions, A_rms_ was 2.43 brpm and 95% LoA were −5.00 to 4.47 brpm (mean bias −0.27 brpm).

**Conclusion:** We showed that the Corsano Cardiowatch 287 using PPG technology can monitor continuous RR and HR with high accuracy in healthy subjects as well as in patients diagnosed with OSA. We conclude that wearable devices like these enable new and more accessible ways to measure health, ultimately improving healthcare delivery. The trial was registered in the ISCRTN registry under reference ISRCTN13965929.

## Introduction

Respiratory rate (RR) is a vital sign which could be abnormal in many pathological, para-physiological and physiological conditions. In adults a normal respiratory rate during rest is generally considered to be between 12 and 20 breaths per second [1]. An abnormal RR could be, in the presence of other suggestive symptoms, supportive of a diagnose in specific cases such as sleep apnea [2], pneumonia [3], sudden infant death syndrome [4,5] and chronic obstructive pulmonary disease [6].

Furthermore, as high RR could be predictive of future critical illness in selected circumstances e.g. at discharge following pulmonary illness [7,8], continuous monitoring could provide clinicians with a real time indicator of their patient’s health. According to the National Institute for Health and Care Excellence [9], RR is the best marker of an ill patient and is the first observation that indicates deterioration in condition.

Current methods for measuring RR include manually counting chest movements, estimated RR from ECG patch devices, spirometry, and capnography [10]. Besides, RR can be measured with respiratory inductance plethysmography (RIP) by measuring the movement of the chest and abdominal wall [11]. RIP is the most frequently used, established and accurate plethysmography method to estimate respiratory movements and has been used in many clinical studies including overnight sleep studies. Unfortunately, all these methods do not allow long term continuous monitoring due to the need for manual supervision, cost or discomfort to the patient.

Liu et al [12] disclose in their topical review of recent developments of RR measurement technologies that portable sensors with wireless communication will play an important role in de future for the early detection of disease and deterioration. The Corsano CardioWatch 287 has been developed in order to provide clinicians with a monitoring device that is able to deliver this. The Corsano CardioWatch 287 is a non-invasive, continuous, wrist-worn and wireless monitoring device based on photoplethysmography (PPG) technology. This enables accessible long term continuous monitoring of RR in ambulatory patients.

Numerous studies have shown that RR can be accurately measured from PPG [13,14]. The plethysmogram contains a direct current (DC) part, representing underlying tissue and venous blood content, and an alternating current (AC) part, reflecting arterial pulsations [15]. Respiration leads to variations in peripheral blood volume, which translate to variations in the DC part of the plethysmogram. Subsequently, RR can be measured by computing the frequency of these variations [16].

Most studies on the accuracy of RR measurements by PPG involve healthy participants in ideal conditions. Meanwhile, several medical conditions affect the plethysmogram in a way that makes RR measurement more challenging. With regard to Obstructive Sleep Apnea (OSA), it has been demonstrated that arousals introduce significant pulse wave amplitude variations, among other changes [17]. This demonstrates the urge for the validating PPG measurements in a more diverse study population.

Several PPG-devices have demonstrated the ability to track sleep and diagnose OSA with acceptable sensitivity but with poor specificity and are therefore unsuitable for clinical settings [18]. This shows the urge to improve clinically oriented PPG-devices to guarantee accurate measurements of RR and HR.

The primary aim of this study is to evaluate the accuracy of the Corsano CardioWatch 287 in measuring respiratory rate and heart rate at rest in a study population containing both healthy participants and patients diagnosed with Obstructive Sleep Apnea (OSA), compared with a gold standard RR reference device. The secondary aims are to assess measurement availability during rest and to evaluate the accuracy of the Corsano CardioWatch 287 under free-living conditions.

## Materials and Methods

### Study design and patient selection

This is a single-center, cross-sectional study assessing the accuracy of the respiratory rate and the heart rate measured by the Corsano CardioWatch 287-1. The study cohort consisted of 8 healthy subjects and 18 patients with suspected Obstructive Sleep Apnea. All patients showed symptoms associated with OSA, screened positive for OSA with the Philips Questionnaire [19] and showed nocturnal desaturation (ODI 2%>12.2) measured overnight with the WristOx 3100 oximeter [20].

All patients were referred to the medical practice for an overnight home sleep apnea testing (HSAT). They were scheduled for this overnight investigation independently of the proposed study. Healthy individuals, not suspected for having OSA, also underwent overnight HSAT. All participants that were scheduled for these examinations, met the inclusion criteria (≥ 18 years old) and underwent simultaneous continuous overnight PPG with the Corsano CardioWatch 287 and HSAT. Furthermore, they all provided written informed consent for participation in the study, which was approved by the ethics committee of Amsterdam University Medical Centra.

Patients who wore a cardiac implanted electronic device at the time of this study, who were unable to wear the PPG-device, who suffered significant mental of cognitive impairment and patients who were enrolled in another clinical investigation in which the intervention might compromise the safety of the subject’s participating in the study, were excluded from participating in this study.

In order to show the device’s accuracy in different conditions and different situations, the study protocol included participants with different skin types, different hair densities and different severity of OSA.

### Corsano CardioWatch 287

The Corsano CardioWatch 287 is a wrist-worn cardiac arrhythmia screening bracelet that can help identify cardiovascular risks in an early phase [21]. Furthermore, the Corsano CardioWatch 287 is intended for use on general care patients as a patient monitor in home or healthcare setting, providing continuous collection of physiological data, including heart rate, heart rate variability, respiratory rate, activity and sleep [21, 22].

The system consists of a CardioWatch 287 measuring device which sends the collected data via Bluetooth Low Energy (BLE) connection to the CardioWatch App on the wearer’s smartphone. From there, the data is transmitted to the CardioWatch cloud for storage and analysis. The device comprises an accelerometer, which provides data on the activity level of the wearer, and a photoplethysmogram (PPG) sensor. The PPG sensor consists of a LED and a photodiode, enabling the measurement of light reflected from the arteries and arterioles in the subcutaneous tissue of the wearer. Similar to the arterial blood pressure waveform, the resulting PPG waveform shows pulsations reflecting heart beats. Additionally, the baseline of the waveform shows a slow modulation at the rate of the respiratory rate. This modulation is caused by a changing venous return due to changes in intra-thoracic pressure during breathing [23]. Proprietary algorithms of Corsano Health exploit these waveform characteristics to determine heart rate and respiratory rate.

### Home sleep apnea testing

In order to get a large range of respiratory rates and heart rates at rest, we included subjects with and without diagnosed Obstructive Sleep Apnea (OSA) and studied them while sleeping. Patients with OSA suffer from complete or partial cessations of breathing during their sleep. These events are called apneas and hypopneas, respectively. They are often accompanied with oxygen desaturations, arousals and heart rate changes. The cessations in airflow will increase the range of respiratory rates.

The severity of OSA can be classified based on the AHI. The AHI is the number of apneas and hypopneas recorded during the study per hour of sleep. It is generally expressed as the number of events per hour. Hypopnea is a 30% or greater decrease of pre-event baseline flow, lasting at least 10 seconds and associated with a 3% or greater oxygen desaturation from pre-event baseline. Apnea is a 90% or greater decrease of pre-events baseline flow, lasting at least 10 seconds [24].

A method for diagnosis of OSA is overnight home sleep apnea testing. Subjects undergo a home sleep study with a validated portable monitoring Noxturnal T3 device from Nox Medical [25]. Measurements include airflow through a nasal cannula, thoracic and abdominal movements, body position, heart rate, activity, oxygen saturation, plethysmograph waveform, snoring and sound.

Thoracic and abdominal movements are measured through respiratory inductance plethysmography (RIP), which requires the wearing of RIP bands around the chest and abdomen. Since inspiration leads to the expansion of chest and abdomen, respiratory rate can be accurately measured by evaluating the time between two maximal band stretches. Additionally, heart rate is measured with a Nonin finger-clip pulse-oximeter (POx) and a Nonin 3150™ WristOx2 Soft Sensor.

### Data acquisition

All participants visited the medical practice where they were prepared for the overnight HSAT and measurements with the PPG bracelet. While at the medical practice, our medical technician placed the recorder with RIP effort bands on the upper body of the subject and placed a cannula for the nose. When the subject would go to sleep that night at home, he/she would consequently place the nasal cannula in the nostrils and put on the wristband with sensor for the pulse-oximeter and the PPG bracelet himself. The pulse-oximeter was placed on the right wrist, while the PPG bracelet was placed on the left wrist. The subject was instructed to tighten the strap from the PPG bracelet that it is secure against the skin; the green sensor light should not be visible and the bracelet should be snug but comfortable. Furthermore, the subject was instructed to wear the bracelet approximately 1 cm below the wrist. The medical technician started the PPG measurements through the app on the mobile device connected to the bracelet. At home, the subject started the HSAT measurements at least half an hour before bedtime in order to obtain free-living data. The next morning, the subject disconnected all medical equipment and returned it to the medical practice. The medical technician verified if the data from the PPG bracelet were transferred to the cloud for storage and downloaded the data from the overnight HSAT to the computer for analysis. The HSAT data were analyzed according to the AASM Manual for the Scoring of Sleep and Associated Events [25] by a trained polysomnographic analyst. The collected respiratory rate and heart rate data were stored for analysis. The quality of data from the PPG bracelet was assessed by a proprietary algorithm of Corsano Health.

The PPG device updated respiratory rate and heart rate every 10 seconds (0.167 Hz). The HSAT device provided 3 Hz respiratory rate data and 20 Hz heart rate data. Both measurement methods used a window of 30 seconds for the computation of respiratory rate.

### Data analysis

We compared each PPG data point with the simultaneously acquired HSAT datapoint. All data points were checked for validity. We excluded all measurements from the analysis that were marked as low quality data or contained unrealistic values. The Corsano CardioWatch makes use of a proprietary quality measure based on the signal amplitude, ranging from 0 (worst) to 4 (best). Data scored with quality index 4 can be considered as free of interference from other sources. For the primary analysis, we included all measurements with a quality index of 4 to assess accuracy in rest conditions. In the secondary analysis, which aims to assess accuracy in free-living conditions, we included all measurements regardless of signal quality to maximize the number of measurement pairs, as signals obtained during motion often have a quality index lower than 4.

The Noxturnal T3 generates unrealistic values when no valid measurement is possible. For example, a failed respiratory rate measurement may yield a default value of −0.015 brpm, and a failed heart rate measurement may yield a default value of either −0.5 or 510 bpm. These values were excluded from the analysis. All HSAT measurements were manually validated by a clinician as part of the routine polygraphic evaluation.

We separated all data into rest and free-living data. The first half hour of the measurements was used for the free-living accuracy analysis. To ensure that the patients did not sleep during this period, the polygraphy data was manually checked. The clinician’s assessment of start and end time of sleep were used to identify measurements obtained under rest conditions.

Considering that we could not guarantee perfect temporal alignment between both measurement devices, we applied a moving average of 60 seconds on both signals. The 60 seconds average was only computed when at least 40 consecutive seconds of valid data was available.

Primary outcome measure was accuracy of PPG for measuring respiratory rate and heart rate compared to HSAT as the gold standard, evaluated for the entire group of subjects, as well as separately for gender and skin color. We assessed accuracy by calculating root-mean-square accuracy

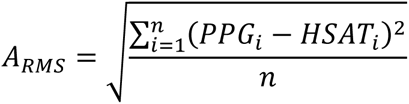

Additionally, we performed Bland Altman Limits of Agreement (LoA) analysis and evaluated the Pearson correlation coefficient between PPG and HSAT.

Bias was calculated as

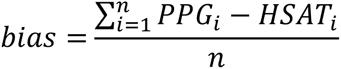

and Limits of Agreement as

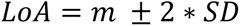

Finally, to assess the influence of patient characteristics on the accuracy of PPG, we conducted a linear regression analysis. Absolute difference of respiratory rate between PPG and HSAT was regarded as the dependent variable, while age, skin color, hair density, BMI and AHI were regarded as independent variables.

Secondary outcome measures were measurement availability, which we defined by the percentage of measurement minutes containing at least 1 valid measurement, and A_rms_ and bias of measuring respiratory rate in free-living conditions.

## Results

### Participant demographics

The study population consisted of 26 participants (participants (47±16 years old, 8 (31%) female). Participants were 18 patients suspected of OSA and 8 healthy subjects. 5 participants were elderly (age>65 years), 6 participants were young (age <30 years) and 11 participants were between 30 and 65 years old. Regarding the BMI, 9 participants were obese (BMI>30), 1 participant was underweight (BMI<18.5) and 16 participants with a BMI between 18,5 and 30. The skin type heterogeneity was limited, as 50% of the participants were classified with a cream white skin (Fitzpatrick type III). 1 subject had dark-toned skin (Fitzpatrick Scale 5 and 6). The hair density (scored as nil, sparse, moderate, dense) varied among the participants, with a predominance of moderate. 2 subjects had a high density of hair in the wrist area where the watch is located. Table 1 displays the baseline characteristics of the participants. 15 of the 18 patients suspected of Obstructive Sleep Apnea tested positive for OSA, with an Apnea Hypopnea Index (AHI) *≥* 5/h. All 8 healthy subjects tested negative for OSA (AHI<5/h). Thus, the study cohort consisted of 11 healthy subjects and 15 patients diagnosed with OSA.

**Table 1.**
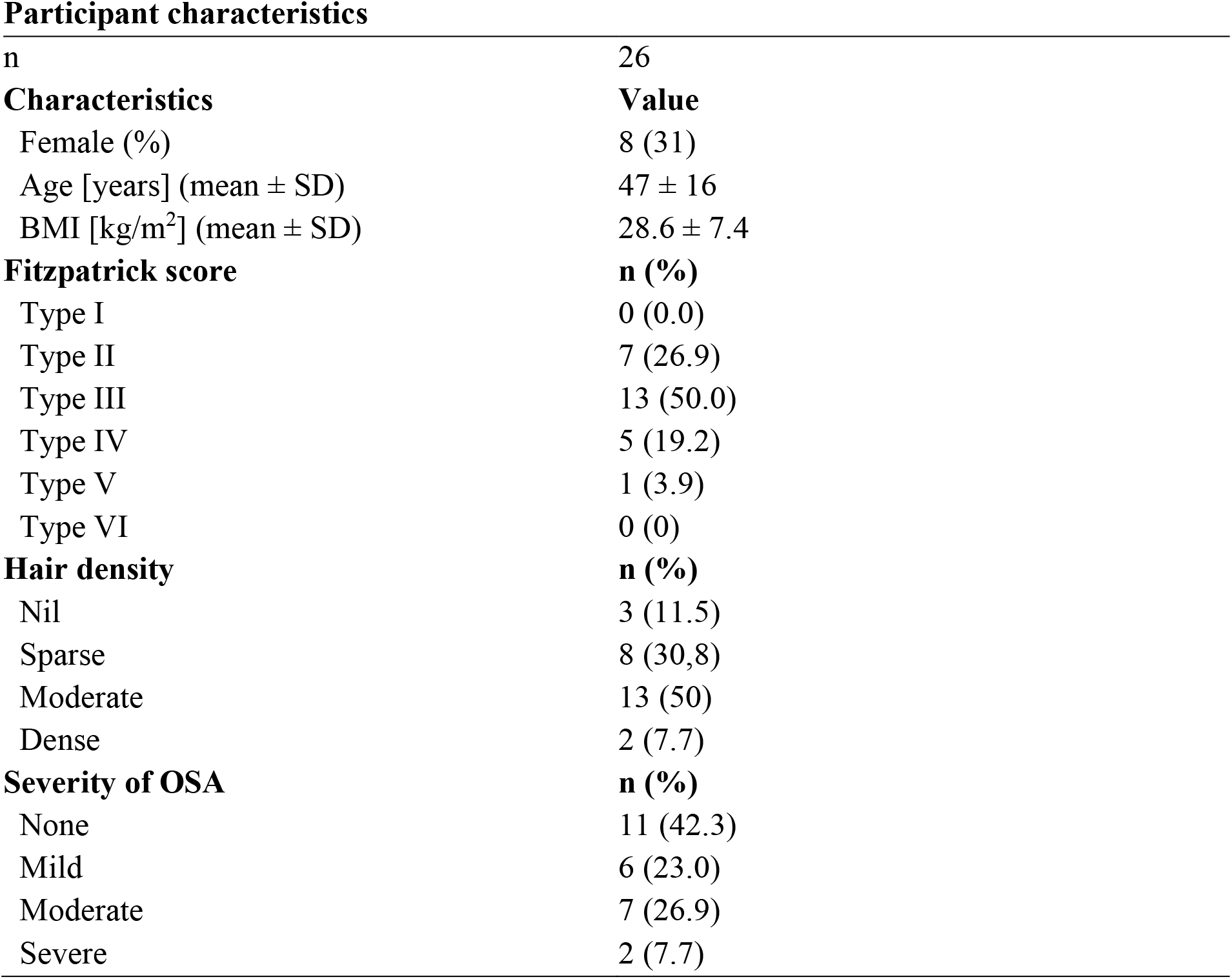
Participant characteristics. *BMI - Body Mass Index. Fitzpatrick scores - type I, pale white; type II, white; type III, cream white; type IV, moderate brown; type V, dark brown; and type VI, deeply pigmented dark brown. Hair density - nil, sparse, moderate and dense. Severity of OSA – none, AHI<5/h; mild, 5≤AHI<15/h; moderate, 15≤AHI<30/h; severe, AHI ≥ 30/h*.

|  |  |
| --- | --- |
| n | 26 |
| **Characteristics** | **Value** |
| Female (%) | 8 (31) |
| Age [years] (mean ± SD) | 47 ± 16 |
| BMI [kg/m<sup>2</sup>] (mean ± SD) | 28.6 ± 7.4 |
| **Fitzpatrick score** | **n (%)** |
| Type I | 0 (0.0) |
| Type II | 7 (26.9) |
| Type III | 13 (50.0) |
| Type IV | 5 (19.2) |
| Type V | 1 (3.9) |
| Type VI | 0 (0) |
| **Hair density** | **n (%)** |
| Nil | 3 (11.5) |
| Sparse | 8 (30.8) |
| Moderate | 13 (50) |
| Dense | 2 (7.7) |
| **Severity of OSA** | **n (%)** |
| None | 11 (42.3) |
| Mild | 6 (23.0) |
| Moderate | 7 (26.9) |
| Severe | 2 (7.7) |

During sleep, a total of 10962 minutes (182 hours and 42 minutes) were simultaneously recorded with HSAT and PPG. As for respiratory rate, 10500 minutes (95.5 %) contained at least one valid PPG measurement, versus 10845 minutes (98.9%) by HSAT. As for heart rate, 10777 minutes (98.3%) contained at least one PPG measurement, versus 10806 minutes (98.6%) by HSAT. Figure 1 shows a typical measurement by both PPG and HSAT.

**Figure 1.**
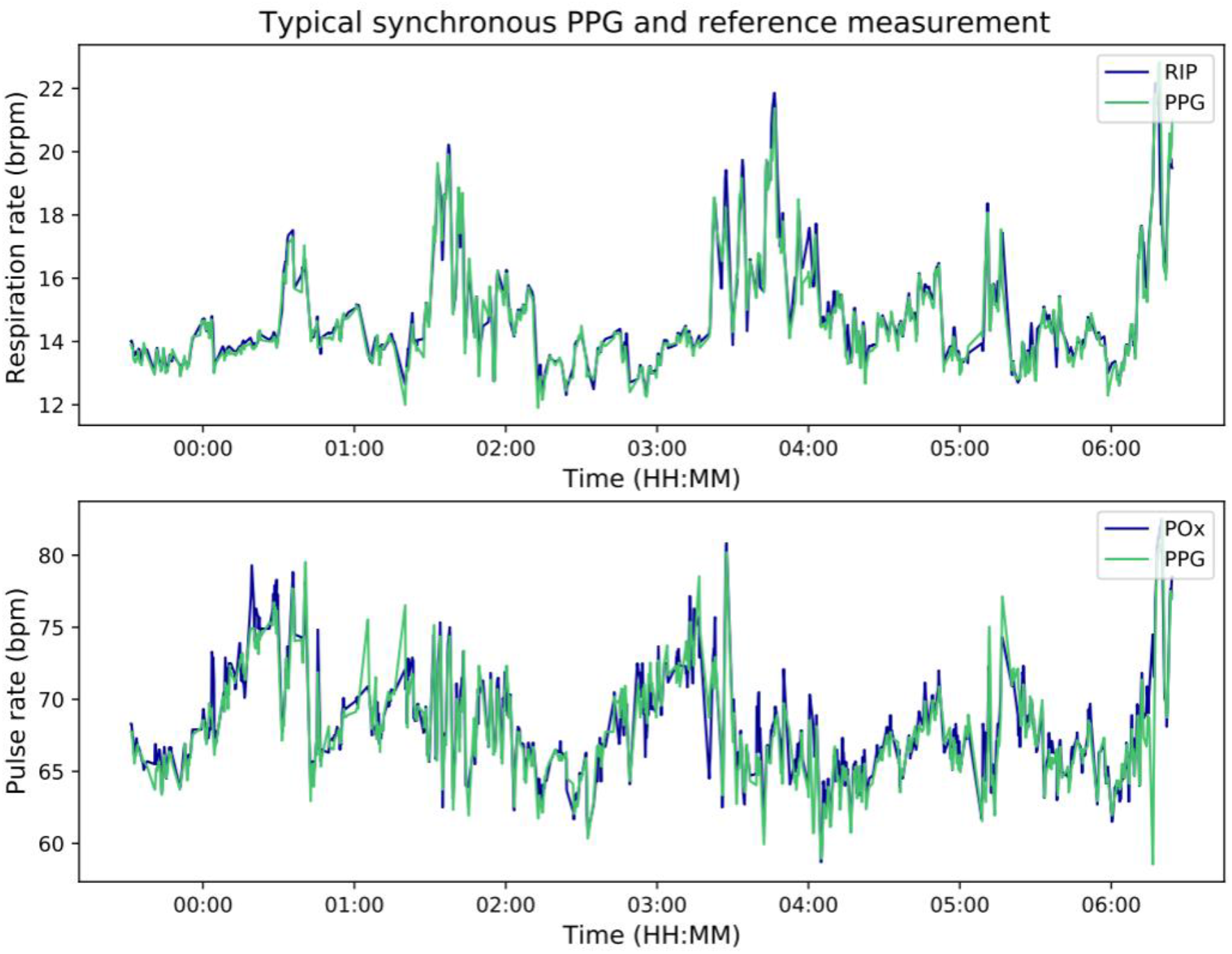
Typical simultaneous PPG and HSAT measurement of respiratory rate (upper) and heart rate (lower). Data is smoothed by a moving average of 60 seconds. *bpm – beats per minute. brpm – breaths per minute. POx – Pulse oximeter. PPG – photoplethysmography. RIP – respiratory inductance plethysmography. HSAT – home sleep apnea testing*.

### Respiratory rate accuracy during rest

In 26 participants, a total of 31083 respiratory rate measurement pairs were available during rest. The PPG A_rms_ was 0.60 brpm. Correlation between PPG and HSAT was high with R = 0.964 (95%CI 0.963 to 0.965) (figure 2). Bias was −0.14 (95%CI −0.21 to −0.08) brpm. The 95% Limits of Agreement were −1.28 (95%CI −1.35 to −1.22) to 1.71 (95%CI 0.93 to 1.06) brpm.

**Figure 2.**
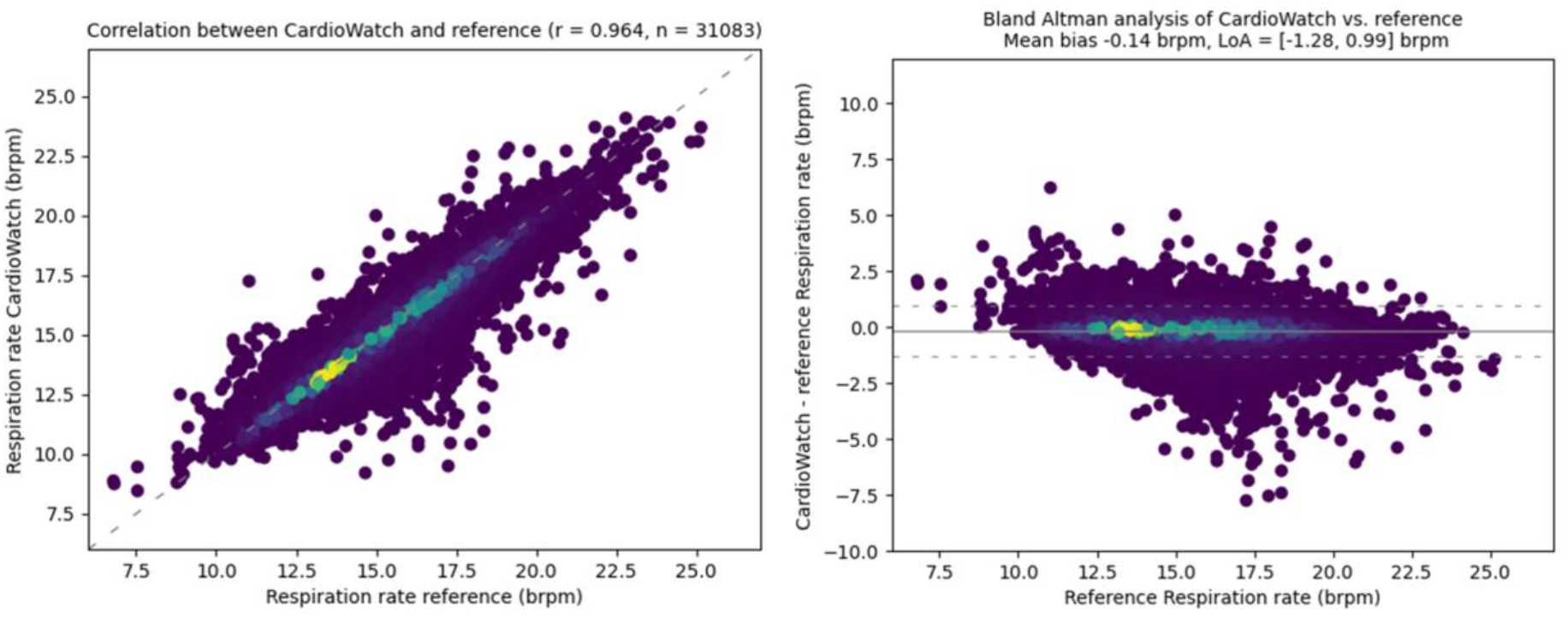
Correlation (left) and Bland Altman (right) analysis of respiratory rate measured by PPG and HSAT. Mean bias was −0.14 (95%CI −0.21 to −0.08) brpm. *brpm – breaths per minute. HSAT – home sleep apnea testing. LoA – Limits of Agreement. PPG – photoplethysmography*.

Table 2 shows the PPG A_rms_ and bias specified for gender and skin color. The supplementary materials include Bland Altman plots for each category.

**Table 2.** PPG respiratory rate accuracy. A_rms_ – root-mean-squared accuracy. brpm – breaths per minute. Fitzpatrick scores – type I, pale white; type II, white; type III, cream white; type IV, moderate brown; type V, dark brown; and type VI, deeply pigmented dark brown

| **Total** | **n** | **A<sub>rms</sub> (brpm)** | **Bias (brpm)** |
| --- | --- | --- | --- |
| Total | 31083 | 0.60 | -0.14 |
| **Gender** | **n** | **A<sub>rms</sub> (brpm)** | **Bias (brpm)** |
| Male | 22124 | 0.65 | -0.16 |
| Female | 8959 | 0.46 | -0.11 |
| **Fitzpatrick score** | **n** | **A<sub>rms</sub> (brpm)** | **Bias (brpm)** |
| Type II | 8342 | 0.46 | -0.15 |
| Type III | 17079 | 0.68 | -0.17 |
| Type IV | 4382 | 0.45 | -0.06 |
| Type V | 1280 | 0.58 | -0.03 |

### Heart rate accuracy during rest

In 26 participants, a total of 38693 heart rate measurement pairs were available during rest. The PPG A_rms_ was 0.95 bpm. Correlation was high with R = 0.996 (95%CI 0.996 to 0.996) (figure 3). Bias was −0.46 (95%CI −0.53 to −0.39) bpm. The 95% Limits of Agreement were −2.09 (95%CI −2.17 to −2.02) to 1.17 (95%CI 1.10 to 1.25) bpm.

**Figure 3.**
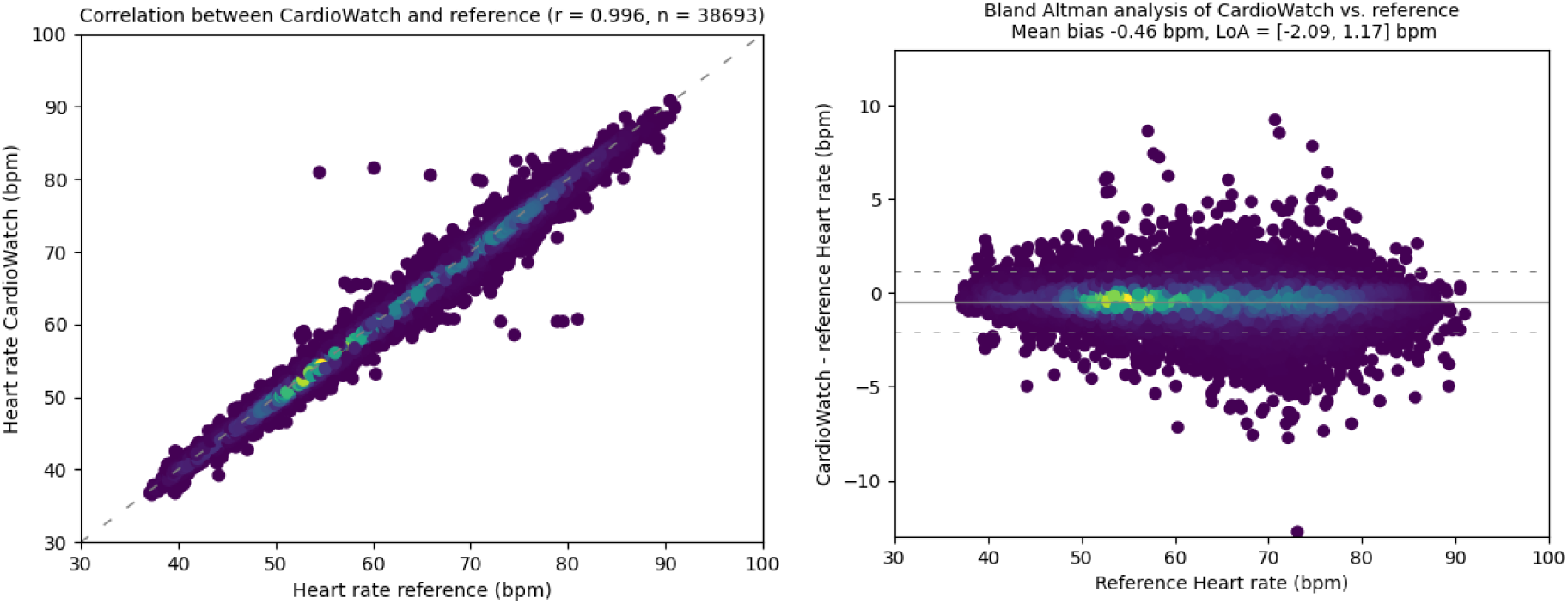
Correlation (left) and Bland Altman (right) analysis of heart rate measured by PPG and HSAT. Mean bias was −0.46 (95%CI −0.53 to −0.39) bpm. *bpm – beats per minute. HSAT – home sleep apnea testing. LoA – Limits of Agreement. PPG – photoplethysmography*.

### Subgroup analysis

Despite low numbers in some groups, a subgroup analysis was performed. This analysis demonstrated low and non-significant coefficients for age, skin colour, hair density, BMI and AHI (table 3).

### Respiratory rate accuracy in free-living conditions

In 26 participants, a total of 4041 respiratory rate measurement pairs were available during free-living conditions. The PPG A_rms_ was 2.43 brpm. Correlation between PPG and HSAT was high with R = 0.801 (95%CI 0.790 to 0.812) (figure 4). Bias was −0.27 (95%CI - 0.34 to −0.19) brpm. The 95% Limits of Agreement were −5.00 (95%CI −5.13 to −4.88) to 4.47 (95%CI 4.34 to 4.59) brpm.

**Figure 4.**
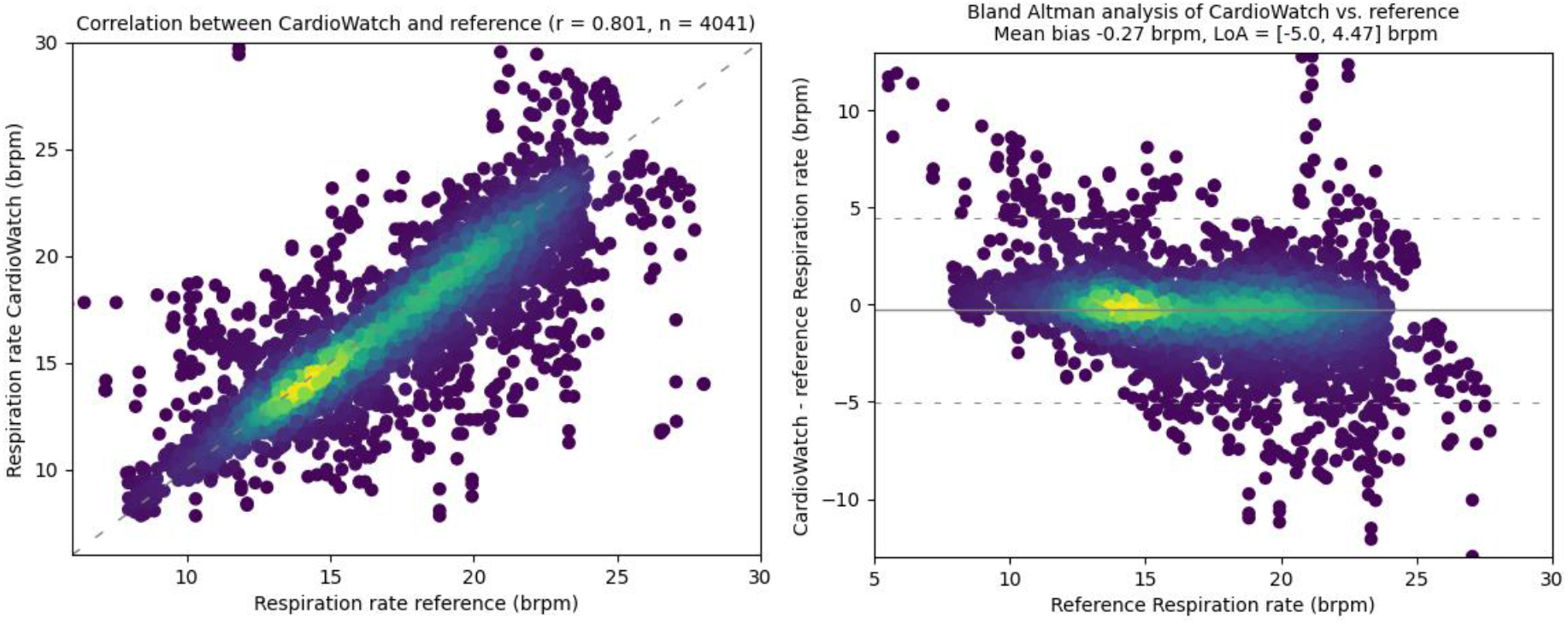
Correlation (left) and Bland Altman (right) analysis of respiratory rate measured by PPG and HSAT in free-living conditions. Mean bias was −0.27 (95%CI −0.34 to −0.19) brpm. *brpm – breaths per minute. HSAT – home sleep apnea testing. LoA – Limits of Agreement. PPG – photoplethysmography*.

## Discussion

### Present Study

We studied the Corsano CardioWatch 287 wristband that uses PPG technology for continuous respiratory and heart rate monitoring and tested its accuracy at rest amongst 26 individuals with varying characteristics. Accuracy was tested against HSAT as gold standard sleep monitoring reference device. We observed accurate respiratory rate and heart rate measurements during rest with > 0.96 correlation between the PPG bracelet and HSAT. PPG measurements were within 1 b(r)pm to reference measurements as quantified by root mean square accuracy, which indicates good accuracy. No significant differences in performance among subgroups were observed. Accuracy in free-living conditions was notably lower but still within 2.5 b(r)pm.

Measuring vital signs like RR and HR enables early detection and monitoring of several conditions and diseases. Continuous home monitoring of RR provides clinicians with an indicator of their patient’s health since abnormal RR is predictive of a future critical illness [7,8] and assists them with the management and monitoring of chronic diseases and postoperative rehabilitation. Furthermore, continuous RR monitoring enables immediate rescue in life-threatening conditions as RR can significantly change in a few minutes. Not only in hospital routines but also in everyday tasks of common people, the monitoring of RR is significant to get an overview of someone’s health.

Continuous HR monitoring is critical to the management of cardiovascular disease as elevated HR is an independent predictor of cardiovascular events, mortality and hospitalization for worsening heart failure [26,27].

The results presented by the current study suggest that the Corsano CardioWatch 287 may be a valuable tool for long-term accurate monitoring of RR and HR.

### Strengths and limitations

A strength of this study is the fact that the study population consists for the bigger part of patients with diagnosed OSA. OSA is characterized by periods of apneas and hypopneas, which is challenging for PPG measurements. After all, while RIP is a very direct method of measuring respiration by measuring expansions of chest and abdomen, PPG signals contain only low powered frequencies representing respiration which are especially difficult to interpret in the extremes. Our study shows that the frequency of hypopneas and apneas, expressed in AHI, does not decrease RR accuracy significantly.

A limitation of this study is that all measurements were done in resting (sleeping) subjects, thus precluding assessment of the device’s ability to discern PPG waveforms from noise, speech and motion artifacts. It is well known that the accuracy of measurements derived from PPG based devices is affected by motion artifacts [28,29]. Further studies must be performed to validate the RR and HR obtained from the PPG waveforms of the CardioWatch 287 in subjects while awake and during different physical activities in order to test the device’s accuracy in these situations.

A second limitation of this study is the fact that the bracelet was evaluated against a pulse-oximeter for assessing heart rate accuracy. Many sleep diagnostic devices contain electrodes that measure the electrocardiogram (EKG), which is the gold standard for monitoring heart rate. Comparing the bracelet’s heart rate accuracy with EKG would be superior. Previous studies have shown that ‘pulse’ rate does not necessarily equates heart rate measured by ECG during periods of heart rate acceleration and deceleration [30]. Patients diagnosed with OSA show repeated episodes of intermittent periods of hypoxia and hypercapnia which lead in turn to a significant increase in sympathetic activation and heart rate variability. Even in healthy subjects the nocturnal heart rate variability depends on the sleep stage; sympathetic nervous system activation during REM sleep causes an increase in heart rate variability. Notwithstanding, we used the pulse oximeter for heart rate comparison. First, our polygraphy device did not contain EKG electrodes and is limited to the use of the pulse oximeter. Considering that in standard clinical practice we make use of pulse oximetry, it is important to know whether the wristband is a good alternative. Secondly, the bracelet’s heart rate accuracy has already been tested against EKG in a different study [21].

Another limitation is that the subjects recruited for the study come with restricted characteristics. No meaningful conclusion could be drawn from the subgroup analysis because of the low numbers in some groups. Despite the good coverage of gender, age, hair density and severity of OSA, the variation in skin tones is limited with a slight skew to the lighter pigmentation range. Furthermore, the range of BMI is limited with predominance of patients with low BMI. Further studies are needed to demonstrate accurate RR and HR measurements in larger populations with more diverse characteristics.

Finally, accuracy of measuring respiratory rate in free-living conditions was tested for a limited period of time. We suggest a future study in which participants are monitored for at least 24 hours to capture a full range of physical activities. However, this will require the use of a high-accuracy, comfortable reference device, which are currently scarce.

### Future role of PPG devices in sleep laboratory

Increasing awareness of the high prevalence of OSA and its impact on health in conjunction with high cost and inconvenience of poly(somno)graphy stimulate the development of more convenient, affordable, and accessible diagnostic devices. Since this study demonstrates a high accuracy of the CardioWatch 287 in measuring respiratory rate and heart rate at rest with no significant decrease in accuracy for patients diagnosed with OSA, wrist-worn PPG devices have the potential to be used in a sleep laboratory for tracking sleep, revealing sleep architecture and possibly detecting OSA. Previous research has shown a good reliability and accuracy of a finger pulse-oximetry PPG based device in diagnosing OSA compared to the golden standard polysomnograhpy [31]. Previous research also shows that wrist-worn PPG devices are very well suited for long-term home sleep monitoring although more evidence is needed to determine whether it can replace poly(somno)graphy with enough accuracy to enable unobtrusive sleep monitoring in clinical practice [22]. Therefore, the role of a wrist-worn PPG-based device in a sleep laboratory looks promising for long-term sleep monitoring but merits more research, especially with a larger clinical population including individuals with sleep disorders.

## Conclusion

This validation study showed that the Corsano Cardiowatch 287 using PPG technology can monitor continuous RR and HR with high accuracy in healthy subjects as well as in patients diagnosed with OSA, with no significant differences in performance among subgroups. Due to their continuous monitoring, noninvasive and convenient-to-patient nature, this device has great potential for becoming part of our everyday lives and act as companion tool for clinical decision support supplementing established gold standard methods. Wearable devices like these enable more accessible ways to measure health in a broader range of healthcare settings, ultimately improving healthcare delivery.

## Informed consent

Informed consent was obtained from all individual participants included in the study.

## Data availability statement

The data that support the findings of this study are available upon reasonable request from the authors.

## Acknowledgement

J.M. Gehring, L.C. Saeijs-van Niel and L.P. ten Bosch-Paniagua contributed to the conception and design of the study and the data acquisition. J.M. Gehring contributed to the analysis and interpretation of the data and drafted the article. L.C. Saeijs-van Niel, L.P. ten Bosch-Paniagua and M.H. Frank critically reviewed and edited the article. L.C. Saeijs-van Niel and L.P. ten Bosch-Paniagua were responsible for the project administration. All authors discussed the results and combined this to the final article and gave approval of the version to be submitted. The authors would like to thank A.R. van Nieuw Amerongen and I.J.G. Hanse for their technical advice and comments.

## Ethical approval

All procedures performed in studies involving human participants were in accordance with the ethical standards of the medical ethical committee of the Amsterdam University Medical Centra (METC AMC) and with the 1964 Helsinki declaration and its later amendments or comparable ethical standards.

## Informed consent

Obtained

